# Pharmacogenomic predictors of drug response and choice in dyslipidemia and hypertension

**DOI:** 10.64898/2026.01.28.26345024

**Authors:** Fumihiko Takeuchi, Malathi S.I. Dona, William W.H. Ho, Samuel A. Lambert, Michael Inouye, Norihiro Kato

**Author notes:** Correspondence Systems Genomics Laboratory, Baker Heart and Diabetes Institute, 75 Commercial Road, Melbourne VIC 3004, Australia.

## Abstract

**Background:** Drug suitability is determined by safety, efficacy, and pathological appropriateness. The pharmacogenomics of drug suitability can be assessed by analyzing drug response and drug choice in large population cohorts.

**Methods:** We investigated drug response and drug choice for dyslipidemia and hypertension using genetic, phenotypic, and prescribing data from the UK Biobank and the All of Us Research Program. Drug response was reassessed with rigorous biomarker scaling, while genome-wide association studies (GWAS) and polygenic scores were used to examine genetic factors influencing drug choice.

**Results:** Conventional analyses showed that variants influencing baseline LDL cholesterol (LDL-C) were inversely associated with absolute LDL-C change but concordant with relative change following statin therapy; these signals disappeared after applying a variance-stabilizing Box–Cox transformation, indicating a methodological artifact in biomarker scaling. GWAS for drug choice identified several significant loci and unique genetic correlation patterns with cardiometabolic traits. Polygenic scores for drug choice yielded statistically significant predictive performance, which was enhanced by incorporating demographic factors, though prediction strength in clinical settings remains modest.

**Conclusion:** Variance-stabilizing transformation corrects spurious pharmacogenetic associations introduced by biomarker scaling. Genetic variation informs drug choice for dyslipidemia and hypertension, but current polygenic scores provide only modest benefits in clinical application.

## 1. Introduction

Drug suitability for a patient depends on safety, efficacy, and pathological appropriateness. This study focuses on the pharmacogenomic determinants of efficacy and overall suitability by investigating drug response and drug choice for dyslipidemia and hypertension in the UK Biobank and the All of Us Research Program.

Pharmacogenomics has characterized the genetic determinants of drug efficacy, particularly in relation to LDL cholesterol (LDL-C) response to statins. Several variants in *LPA*, *APOE*, *SORT1*, and *SLCO1B1* genes influence statin response [1,2], and most of these variants are also associated with baseline LDL-C levels [3]. In contrast, genome-wide association studies (GWAS) on blood pressure response to antihypertensive medications have not yet identified any loci reaching genome-wide significance [4,5].

In replicating prior GWAS on statin response, we analyze both absolute and relative LDL-C changes as phenotypes, following conventional methodologies. However, our findings suggest that previously reported associations may be artifacts introduced by the scaling methods: raw scale for absolute change and log-scale for relative change. Notably, when employing a variance-stabilizing transformation, which serves as an intermediate approach between these two scales, the previously observed association signals disappear.

Drug selection refers to the process of choosing specific drugs for a particular patient population based on scientific evaluations of their safety, clinical efficacy, and pathological appropriateness. Outside oncology, drug selection based on genetic factors remains uncommon, as disease subclassification primarily rely on clinical measurements. Optimal drug treatment for cardiometabolic diseases such as dyslipidemia and hypertension requires both the appropriate type (or class) for the target disease and the appropriate dosage for the individual patient. Cholesterol medications, including statin and ezetimibe for lowering LDL-C and previously fibrate for lowering triglycerides, are prescribed when cardiovascular risk becomes elevated (https://www.nice.org.uk/guidance/ng238). For hypertension, first-line treatment typically includes diuretics, calcium channel blockers or renin-angiotensin system (RAS)-acting agents (https://www.nice.org.uk/guidance/NG136, https://www.who.int/publications/i/item/9789240033986). If blood pressure remains uncontrolled, dose escalation or addition of β-blockers follows. Even when standard medications and dosages are administered according to clinical guidelines, considerable interindividual variation exists in both efficacy and risk of adverse events. The drug selection process considers comorbidities, age, ethnicity and biomarker responses observed during titration, which can extend over several months.

This study aims to integrate genetic information into the drug selection process. We envision a scenario in which a newly diagnosed dyslipidemic or hypertensive patient receives a drug recommendation informed by their genomic profile. Our underlying hypothesis is that prescription patterns in large biobanks reveal genome-prescription associations that can inform future drug choices. The trait of interest is the choice of drug *X* for disease *Y*, defining cases as patients prescribed drug *X* for disease *Y* and controls as disease *Y* patients not prescribed drug *X*. As we utilize real-world prescription data, the observed drug choice reflects the disease severity, comorbidities, and prior drug response. This “drug choice” trait differs from the previously studied “drug use” trait, which defined controls as individuals not prescribed the drug regardless of disease status [6]. We conduct pharmacogenomic analyses of three types of dyslipidemia treatments (statin, fibrate, and ezetimibe) and four classes of antihypertensive drugs (diuretic, β-blocker, calcium channel blocker, and RAS-acting agents) using genetic and phenotypic data collected from large general population cohorts. We examine the genetic basis of drug choice and assess the feasibility of drug recommendation using germline genetic variation by developing polygenic scores (PGS) for drug choice. This drug choice PGS exhibited modest but statistically significant predictability regarding the appropriateness of prescribing specific drug classes to a given patient population.

## 2. Methods

### 2.1. UK Biobank

The UK Biobank is a large-scale biomedical resource with genetic, health, and lifestyle data for approximately 500,000 UK participants enrolled from 2006 to 2010 [7].

For drug efficacy analysis, we used drug prescriptions and laboratory measurements from primary care with records spanning roughly from 2000 to censoring in 2017. Biomarker responses to lipid-modifying drugs (ATC code C10) and antihypertensives (ATC codes C02-C09) were analyzed by comparing baseline and on-drug biomarker levels within a single drug class, excluding periods when other drugs for the same disease were prescribed. For the statin analysis, periods with concurrent use of fibrates or ezetimibe were excluded. Baseline measurements follow a drug-free period of ≥180 days and precede medication initiation by ≤730 days, while on-drug measurements occur 28–730 days post-initiation. The 28-day period marks the time for full therapeutic effect [8,9]. When multiple measurements exist within either period, we calculated linearly weighted averages that gave higher weight to measurements closer to the drug initiation. Prescription episodes were defined as continuous series of prescriptions for the same drug class, allowing ≤14 days gap between prescriptions. The coding for prescriptions and measurements are listed in https://github.com/fumi-github/drug_selection/tree/master/ukb/drugresponse

For drug choice analysis, medication data were obtained from nurse-led verbal interviews during the initial assessment (2006-2010, Data-Field 20003), and drug names were mapped to ATC codes[6]. For omega-3-triglycerides (C10AX06), only prescription Omacor 1g capsules were included, excluding dietary supplements.

Subjects were considered dyslipidemic if they had a diagnosis or self-reported dyslipidemia (ICD-10 code E78) by the initial assessment date, or if their LDL-C ≥190 mg/dL or HDL-C <40 mg/dL at initial assessment [10]. Similarly, subjects were classified as hypertensive if they had a diagnosis or self-reported hypertension (ICD-10 codes I10-I15) or recorded DBP ≥100 mmHg or SBP ≥160 mmHg [11].

Analyses were restricted to self-identified White British participants with consistent genetic ancestry [12], labeled as EUR (European descent), and participants of Indian, Pakistani, or Bangladeshi background by self-identification and genetic ancestry, labeled as SAS (South Asian descent). Genotypes were assayed using Affymetrix microarrays and imputed to the Haplotype Reference Consortium and UK10K reference panels (Data Category 100319) [12]. PGS for LDL-C and hypertension were obtained from Data-Field 26250 and 26244, respectively.

### 2.2. All of Us Research Program

The All of Us Research Program is a diverse cohort of approximately 630,000 USA participants enrolled since 2017 [13]. This study used data from the All of Us Research Program’s Controlled Tier Dataset v8, available to authorized users on the Research Workbench (workspace aou-rw-f61c46ea).

Drug prescription and laboratory measurement data were derived from electronic health records spanning roughly from 2010 to censoring in 2023. Drug efficacy analysis followed the same procedure as for UK Biobank, while drug choice was defined by prescription records extending beyond three months. Dyslipidemia was defined using the condition “Disorder of lipoprotein AND/OR lipid metabolism” (SNOMED code 48286001), and hypertension using “Hypertensive disorder” (SNOMED code 38341003). Analyses were restricted to participants with genetically predicted European descent. Genotyping was performed by short-read whole genome sequencing [14].

### 2.3. Variance-stabilizing Box-Cox transformation

The Box-Cox transformation is a family of power transformation that includes both the identity transformation (parameter λ = 1) and the logarithmic transformation (λ = 0) as special cases. We introduced variance-stabilizing Box-Cox transformation to equalize variances between baseline biomarker level *X* and on-drug level *Y*. The optimal transformation parameter was identified by minimizing the absolute value of the logarithm of the variance ratio, |log(Var [*Y*′] ⁄ Var [*X*′])|, for the transformed baseline and on-drug levels *X’* and *Y’*. The optimal parameter depends on the biomarker-drug combination and the cohort characteristics.

### 2.4. GWAS

To identify loci associated with drug response and drug choice, we conducted GWAS using Regenie software (version 3.4.1.gz) [15]. The analysis involved two steps: computing per-chromosome genetic predictions for SNPs directly genotyped on microarrays or a subset of mutually independent variants (for All of Us, plink2--indep-pairwise 200kb 1 0.5), followed by association testing with all SNPs. In Step 1, SNPs were excluded if they had minor allele frequency (MAF) <0.01, minor allele count <100, missing call rate >0.1, or Hardy-Weinberg equilibrium P-value <10^−15^. Subjects with missing call rates >0.1 were removed. In Step 2, multiallelic SNPs and those with MAF <0.01 were excluded. Additional filters removed imputed SNPs with imputation accuracy ≤0.4, and sequenced SNPs with missing call rate >0.1 or Hardy-Weinberg deviation beyond ±20%. For binary traits, Firth likelihood ratio tests were applied to variants with P-value <0.01.

For the drug choice analysis, covariates included sex, age, BMI, age^2^, sex-by-age interaction, and genetic principal components (top 20 for UK Biobank and top 16 for All of Us). The resulting linkage disequilibrium (LD) score regression intercepts (0.999–1.039 for dyslipidemia drug choice and 1.018–1.036 for hypertension drug choice) indicate absence of population stratification.

For the drug response analysis, to avoid improper adjustment of the composite phenotype (the difference between baseline and on-drug biomarker levels), only genetic principal components were used as covariates. GWAS summary statistics from UK Biobank and All of Us were combined via meta-analysis using the METAL software (version 2020-05-05) [16], limited to SNPs present in both datasets. The resulting LD score regression intercepts (0.989–0.994 for LDL-C response and 0.897–0.994 for SBP response) likewise indicate negligible population stratification. Manhattan plots were drawn using the qqman package (version 0.1.9) [17] of the R software (version 4.4.0) [https://cran.r-project.org/].

Genome-wide significance was defined as P-value <5×10^−8^. Significant SNPs were compared against previous GWAS results using the hugeamp database [https://hugeamp.org/] and the GWAS ATLAS [https://atlas.ctglab.nl/PheWAS].

Our drug choice GWAS was compared with the drug use GWAS from FinnGen [18], in which cases were defined as individuals who purchased the drug and controls as all other participants in the cohort [https://r12.finngen.fi/pheno/RX_STATIN https://risteys.finngen.fi/endpoints/RX_STATIN https://r12.finngen.fi/pheno/RX_ANTIHYP https://risteys.finngen.fi/endpoints/RX_ANTIHYP].

### 2.5. Heritability and genetic correlation

Because drug choice is a binary trait (prescribed vs. not prescribed) defined among patients with a given disease, we estimated heritability specific to the patient subgroup, quantifying the proportion of drug choice variance explained by SNPs. Heritability and genetic correlations were estimated using LD score regression (LDSC software version 1.0.1) [19], which was previously applied in a sex-specific heritability study [20]. Input data included GWAS summary statistics recomputed using PLINK software (version 2.00a2.3LM) [https://www.cog-genomics.org/plink/2.0/] under similar quality control criteria and covariates (excluding the sex-by-age interaction term), published GWAS summary statistics, and the HapMap3 European dataset as reference. Correlation matrices were visualized using the corrplot package (version 0.94) of the R software.

### 2.6. Development of drug choice PGS

To predict drug choice traits, we developed PGS by combining index SNPs from external GWAS for relevant disease traits and fitting weights. We took this approach of weighting the index SNPs, rather than developing PGS from scratch based on drug choice GWAS, because of the limited sample size of the GWAS and to leverage the genetic correlation between drug choice traits and disease traits. An individual’s PGS was calculated as a weighted sum of allele-dose genotypes (coded 0, 1, or 2). To ensure proper validation, index SNPs were taken from disease GWAS that excluded UK Biobank and All of Us, namely studies on lipids [21], blood pressure [22–24], type 2 diabetes [25], BMI [26], and CAD [27]. After excluding SNPs with MAF <0.01 or located on sex chromosomes, 570 SNPs remained.

A basic score was defined as a weighted sum of sex, age, and BMI, while a combined score summed the basic score and PGS. Development of these scores was performed in R using the glmnet package (version 4.1-7). The UK Biobank EUR subjects were split into 80% for model training and tuning and 20% for blind testing (in 20 times repeated independent sub-sampling). Within the 80% subset, data was further divided into 60% for training and 20% for tuning (repeated 20 times) to fine-tune the Lasso penalty hyperparameter.

On the training set, we first performed logistic regression with drug choice as the dependent variable and demographic factors (sex, age, and BMI) as independent variables, including 20 genetic principal components as covariates. The resulting coefficients defined the weights of the basic score. Next, we took the residuals from this regression and applied Lasso regressions (penalty chosen from 0.001 to 1) with SNPs as independent variables. The resulting coefficients defined the weights of a tentative PGS. The variable selection functionality of Lasso regression assigns zero weights to SNPs in LD with other SNPs that show stronger association.

On the tuning set, we performed logistic regressions with drug choice as the dependent variable and the tentative PGS (trained under various penalties) as the independent variable, and selected the Lasso penalty yielding the highest Z score as the optimal hyperparameter. Finally, to scale both scores accurately, we ran another logistic regression incorporating the basic score and the tentative PGS as independent variables. The coefficient for the basic score was approximately one because we are essentially replicating the logistic regression computed on the training set, indicating that no further rescaling was needed. By contrast, the coefficient for the tentative PGS, this time in an unpenalized regression and concurrently modeled with the basic score which is a stronger predictor, could differ from one and was defined as the scaling factor. We obtained the final PGS by multiplying the tentative PGS weights by this scaling factor. As a result, both the basic score and the final PGS were scaled to have a unit effect size on the log-odds of drug choice.

### 2.7. Validation of drug choice PGS

Internal validation used the reserved 20% of UK Biobank EUR subjects. External validation was performed using the All of Us cohort and the UK Biobank SAS.

Predictive performance was evaluated using the Area Under the Receiver Operating Characteristic Curve (AUC-ROC) analyses for three models: PGS alone, basic score alone, and their sum. Additionally, we evaluated prediction accuracy by analyzing drug choice frequency across score deciles for each model.

For external validation, the final SNP weights were derived by averaging across all 20 cross-validation runs in the UK Biobank EUR. SNP weight signs were corrected for differences in genome builds (GRCh37 in UK Biobank and GRCh38 in All of Us) using the triple-liftOver program (version 1.33) [28].

## 3. Results

### 3.1. GWAS for drug efficacy

We analyzed LDL-C response to hypolipidemic drugs and systolic blood pressure (SBP) response to antihypertensive drugs using data from the UK Biobank and the All of Us Research Program (Table 1). We performed GWAS for medication-biomarker combinations with >1000 observations in both cohorts. For those combinations, on-drug biomarker levels were significantly lower than baseline levels in all except for SBP response to calcium channel blockers in All of Us.

**Table 1A.** Characteristics of subjects analyzed for hypolipidemic response.

|  | UK Biobank, EUR |  | All of Us, EUR |  |
| --- | --- | --- | --- | --- |
| Total subjects | 15,426 |  | 14,894 |  |
| Sex male (N, %) | 8,518 | 55% | 7,488 | 51% |
| Age (mean, SD) | 61.3 | 6.0 | 69.4 | 11.0 |
| BMI (mean, SD) | 28.7 | 4.8 | 30.2 | 6.3 |
| C10AA, Statin |  |  |  |  |
| Drug response observed (N) | 15,217 |  | 13,775 |  |
| LDL-C baseline [mmol/L] (mean, SD) | 4.04 | 0.96 | 3.20 | 1.01 |
| LDL-C on-drug [mmol/L] (mean, SD) | 2.30 | 0.69 * | 2.32 | 0.81 * |
| C10AB, Fibrate |  |  |  |  |
| Drug response observed (N) | 78 |  | 404 |  |
| LDL-C baseline [mmol/L] (mean, SD) | 4.20 | 1.11 | 2.75 | 0.88 |
| LDL-C on-drug [mmol/L] (mean, SD) | 3.56 | 0.92 * | 2.89 | 0.93 |
| C10AC, Bile acid sequestrants |  |  |  |  |
| Drug response observed (N) | 28 |  | 180 |  |
| LDL-C baseline [mmol/L] (mean, SD) | 3.44 | 1.12 | 2.95 | 0.95 |
| LDL-C on-drug [mmol/L] (mean, SD) | 3.05 | 1.12 | 2.81 | 0.90 |
| C10AD, Nicotinic acid and derivatives |  |  |  |  |
| Drug response observed (N) | 4 |  | 313 |  |
| LDL-C baseline [mmol/L] (mean, SD) | 4.79 | 0.63 | 3.07 | 0.91 |
| LDL-C on-drug [mmol/L] (mean, SD) | 4.38 | 0.62 | 3.02 | 0.94 |
| C10AX06, Omega-3-triglycerides |  |  |  |  |
| Drug response observed (N) | 23 |  | 258 |  |
| LDL-C baseline [mmol/L] (mean, SD) | 3.15 | 1.06 | 3.03 | 0.84 |
| LDL-C on-drug [mmol/L] (mean, SD) | 3.45 | 0.96 | 3.10 | 0.85 |
| C10AX09, Ezetimibe |  |  |  |  |
| Drug response observed (N) | 151 |  | 273 |  |
| LDL-C baseline [mmol/L] (mean, SD) | 4.16 | 0.93 | 3.61 | 1.08 |
| LDL-C on-drug [mmol/L] (mean, SD) | 3.27 | 0.81 * | 3.02 | 0.97 * |

Several variants have previously been reported as genetic predictors of LDL-C response to statins [1,2]. We quantified LDL-C response using two conventional metrics: absolute change (on-drug level *Y* minus baseline level *X*) and relative change (log(*Y*/*X*)). When combining results from the UK Biobank and All of Us, we identified genome-wide significant associations at the *APOE*, *LDLR* and *LPA* loci, consistent with previous reports (Figure 1A).

**Figure 1.**
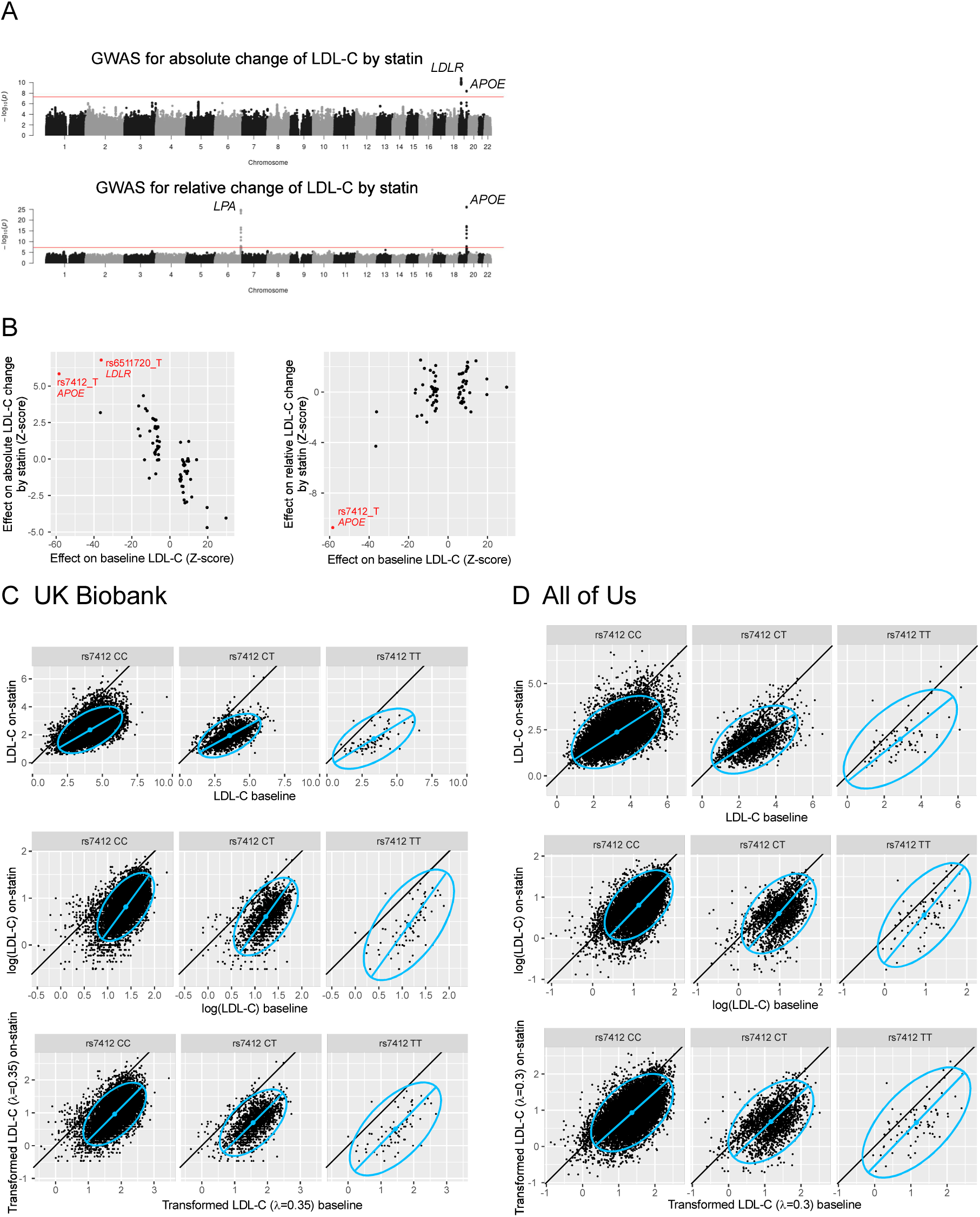
GWAS for LDL-C response on statin. **A**. Manhattan plots of GWAS for LDL-C response to statin based on two phenotype definitions: absolute LDL-C change (top) and relative change (bottom). Variants are plotted as points, with chromosomal position along the horizontal axis and –log_10_(P-value) along the vertical axis. The red line marks the genome-wide significant level of P = 5×10^−8^. **B**. For 71 genetic loci previously associated with baseline LDL-C, the effects of index SNPs on baseline LDL-C (horizontal axis) are compared with their effects on absolute LDL-C change by statin (vertical axis of left panel) and relative change (vertical axis of right panel). SNPs highlighted in red reached genome-wide significance for drug response. In UK Biobank (**C**) and All of Us (**D**), the baseline LDL-C levels of participants (horizontal axis) are compared with the on-statin levels (vertical axis), where LDL-C is scaled in raw scale (top panels), log-scale (middle panels) and variance-stabilizing Box-Cox scale (bottom panels). Points represent individual participants, separated by genotypes of SNP rs7412. Blue ellipses indicate the 95-percentile contours of a fitted bivariate normal distributions, and the blue line segments shows the first principal component, visualizing the data distribution. Black diagonal lines mark equality between the two axes.

The *APOE* locus demonstrated associations with both absolute and relative LDL-C change in response to statin therapy. However, we observed a paradoxical pattern where the T allele of SNP rs7412, which encodes the *APOE* E2 isoform and is associated with reduced baseline LDL-C, exhibited a positive effect on absolute LDL-C change (beta = 0.080, P = 4.8×10^−9^) but a negative effect on relative change (beta = –0.051, P = 6.6×10^−27^). This contradictory pattern was not due to analytical error, as similar results were observed in a separate UK Biobank study [29], where the same allele a showed positive effect on absolute change (beta = 0.053, P = 0.011) and a negative effect on relative change (beta = –0.232, P = 1.1×10^−28^).

To assess whether this pattern generalizes, we systematically examined 71 genetic loci previously associated with baseline LDL-C in an independent GWAS [21] that did not include UK Biobank or All of Us participants. Variants’ effects on baseline LDL-C were systematically opposite to their effects on absolute LDL-C change (Figure 1B, left panel) but aligned with their effects on relative LDL-C change (Figure 1B, right panel). This consistent inversion pattern suggests a methodological artifact rather than genuine biological effects on drug response.

### 3.2. Scaling of biomarker levels

To explore the mechanism underlying this consistent inversion, we examined the joint distribution of baseline and on-statin LDL-C values. In UK Biobank, on-statin LDL-C levels were consistently lower than baseline levels, but the magnitude of reduction was smaller in subjects with lower baseline LDL-C (Figure 1C, top panel). This relationship is illustrated by the distribution-fitting blue line with a slope less than 45 degrees, which was consistent across different genotypes of rs7412. Subjects with the TT genotype, who have lower baseline LDL-C, experienced less absolute decreases, explaining the positive association between rs7412_T and absolute LDL-C change.

For relative change analysis, we examined the distribution after logarithmic transformation of both baseline and on-statin LDL-C values (Figure 1C, middle panel). Under log-scale, subjects with lower baseline LDL-C showed proportionally larger reductions, resulting in subjects with TT genotype experiencing greater relative decreases, which explains the negative association between rs7412_T and relative LDL-C change. The same patterns were observed in All of Us data (Figure 1D, top and middle panels).

This phenomenon arises from unequal variance between baseline and on-statin LDL-C distributions. Pharmacogenetic studies of drug efficacy aim to identify genetic variants influencing the change from baseline (*X*) to on-drug (*Y*) levels while excluding variants associated solely with either *X* or *Y*. However, this goal is compromised when the change measure (*Y* – *X*) is strongly correlated with *X* or *Y*. Under raw scaling (absolute change), the distribution is elongated horizontally, with greater variance in *X* than in *Y*, producing a strong correlation between *Y* − *X* (≈ −*X*) and *X* (r = –0.70 in UK Biobank; Figure 1C, top panel). Under logarithmic transformation (relative change), the distribution elongates vertically with greater variance in *Y* than *X*, yielding a strong correlation between *Y* − *X* (≈ *Y*) and *Y* (r = 0.60 in UK Biobank; Figure 1C, middle panel).

Both raw scaling and log-scaling are special cases of the Box-Cox family of transformations with parameters λ = 1 and 0, respectively. To address these artifacts, we applied a variance-stabilizing Box-Cox transformation to equalize the variances between baseline and on-drug biomarker levels. Under this transformation, the distribution axis approaches a 45-degree angle, minimizing spurious correlations. The optimal Box-Cox parameters were λ = 0.35 for UK Biobank (Figure 1C, bottom panel) and λ = 0.3 for All of Us (Figure 1D, bottom panel). The optimal λ values for SBP response on diuretic, β-blocker, calcium channel blocker and RAS-acting agents were 2.00, –1.15, –1.15, –0.20 in UK Biobank and 1.85, 2.00, –0.75, –0.70 in All of Us. In situation of baseline biomarker level being higher than the on-drug level, the optimal λ becomes high when the baseline standard deviation is smaller than the on-drug standard deviation (as for diuretic, see Table 1B), and low when opposite (as for β-blocker, calcium channel blocker in UK Biobank and RAS-acting agents in both cohorts). Importantly, when we performed GWAS using these variance-stabilized measures, no genome-wide significant associations were detected for LDL-C response to hypolipidemics or SBP response to antihypertensives in the combined analysis of UK Biobank and All of Us (Supplementary Figure 1).

**Table 1B.** Characteristics of subjects analyze for antihypertensive response.

|  | UK Biobank, EUR |  | All of Us, EUR |  |
| --- | --- | --- | --- | --- |
| Total subjects | 28,189 |  | 26,926 |  |
| Sex male (N, %) | 14,276 | 51% | 10,482 | 39% |
| Age (mean, SD) | 59.8 | 6.9 | 63.2 | 15.1 |
| BMI (mean, SD) | 28.9 | 5.0 | 30.0 | 6.8 |
| C02, Various antihypertensives |  |  |  |  |
| Drug response observed (N) | - |  | 2,300 |  |
| SBP baseline [mmHg] (mean, SD) | - | - | 125.0 | 13.2 |
| SBP on-drug [mmHg] (mean, SD) | - | - | 124.8 | 13.7 |
| C03, Diuretic |  |  |  |  |
| Drug response observed (N) | 5,269 |  | 6,230 |  |
| SBP baseline [mmHg] (mean, SD) | 158.3 | 13.8 | 123.9 | 13.1 |
| SBP on-drug [mmHg] (mean, SD) | 146.3 | 14.6 * | 123.1 | 13.8 * |
| C07, β-blocker | | | | |
| Drug response observed (N) | 5,913 |  | 11,403 |  |
| SBP baseline [mmHg] (mean, SD) | 144.8 | 19.1 | 125.9 | 13.7 |
| SBP on-drug [mmHg] (mean, SD) | 135.2 | 16.8 * | 124.8 | 13.8 * |
| C08, Calcium channel blocker |  |  |  |  |
| Drug response observed (N) | 7,428 |  | 1,858 |  |
| SBP baseline [mmHg] (mean, SD) | 156.4 | 15.6 | 124.3 | 12.4 |
| SBP on-drug [mmHg] (mean, SD) | 141.3 | 13.3 * | 124.6 | 13.0 |
| C09, RAS-acting agents |  |  |  |  |
| Drug response observed (N) | 10,723 |  | 7,498 |  |
| SBP baseline [mmHg] (mean, SD) | 154.0 | 15.6 | 135.9 | 14.4 |
| SBP on-drug [mmHg] (mean, SD) | 140.6 | 14.5 * | 131.1 | 13.5 * |

ATC codes of drugs are indicated. EUR, European descent
\* On-drug biomarker level is lower than baseline ( $P < 0.05$ ).

### 3.3. GWAS for drug choice

We next explored the genetic architecture underlying drug choice among patients with dyslipidemia or hypertension by comparing those prescribed a particular drug with those who were not. In UK Biobank European descent (EUR) subjects with dyslipidemia (Table 2A), GWAS on drug choice identified multiple significant loci: 12 for statin, 3 for fibrate, and 2 for ezetimibe (Table 3). These loci overlapped with previously reported associations for lipid traits as well as blood pressure, coronary artery disease (CAD), and type 2 diabetes (T2D). Comparison with statin *use* GWAS in FinnGen [18] showed replication for 13 of 14 SNPs (93%) at genome-wide significance.

**Table 2A.** Characteristics and drug usage of the dyslipidemia cohort.

|  | UK Biobank, EUR |  | All of Us, EUR |  | UK Biobank, SAS |  |
| --- | --- | --- | --- | --- | --- | --- |
| Total subjects | 116,305 |  | 87,006 |  | 3,367 |  |
| Sex male (N, %) | 69,587 | 60% | 39,274 | 46% | 2,269 | 67% |
| Age (mean, SD) | 59.0 | 7.4 | 63.9 | 12.9 | 54.7 | 8.5 |
| BMI (mean, SD) | 28.9 | 4.6 | 30.2 | 6.6 | 28.0 | 4.2 |
| Diabetes (N, %) | 7,535 | 6% | 25,997 | 30% | 584 | 17% |
| Hypertension (N, %) | 58,323 | 50% | 61,159 | 70% | 1,580 | 47% |
| Ischemic heart diseases (N, %) | 15,658 | 13% | 16,615 | 19% | 592 | 18% |
| Drug usage (N, %) |  |  |  |  |  |  |
| C10AA, Statin | 53,671 | 46% | 45,875 | 53% | 1,679 | 50% |
| C10AB, Fibrate | 1,113 | 1% | 3,697 | 4% | 41 | 1% |
| C10AC, Bile acid sequestrants | 97 | 0% | 1,916 | 2% | 0 | 0% |
| C10AD, Nicotinic acid and derivatives | 74 | 0% | 2,856 | 3% | 3 | 0% |
| C10AX06, Omega-3-triglycerides | 404 | 0% | 2,844 | 3% | 21 | 1% |
| C10AX09, Ezetimibe | 2,666 | 2% | 4,162 | 5% | 74 | 2% |

**Table 2B.** Characteristics and drug usage of the hypertension cohort.

|  | UK Biobank, EUR |  | All of Us, EUR |  | UK Biobank, SAS |  |
| --- | --- | --- | --- | --- | --- | --- |
| Total subjects | 137,348 |  | 80,447 |  | 2,896 |  |
| Sex male (N, %) | 73,035 | 53% | 36,892 | 46% | 1,695 | 59% |
| Age (mean, SD) | 60.0 | 6.9 | 63.5 | 13.7 | 57.2 | 7.9 |
| BMI (mean, SD) | 29.0 | 4.8 | 31.0 | 6.9 | 28.4 | 4.5 |
| Diabetes (N, %) | 8,159 | 6% | 26,255 | 33% | 564 | 19% |
| Dyslipidaemia (N, %) | 58,323 | 42% | 61,159 | 76% | 1,580 | 55% |
| Ischemic heart diseases (N, %) | 14,625 | 11% | 16,555 | 21% | 571 | 20% |
| Drug usage (N, %) |  |  |  |  |  |  |
| C02, Various antihypertensives | 5,249 | 4% | 13,370 | 17% | 126 | 4% |
| C03, Diuretic | 29,764 | 22% | 49,822 | 62% | 489 | 17% |
| C07, β-blocker | 22,369 | 16% | 48,476 | 60% | 533 | 18% |
| C08, Calcium channel blocker | 27,049 | 20% | 27,230 | 34% | 813 | 28% |
| C09, RAS-acting agents | 52,788 | 38% | 47,589 | 59% | 1,363 | 47% |

ATC codes of drugs are indicated. EUR, European descent; SAS, South Asian descent

**Table 3.** Significant loci for GWAS of dyslipidemia drug choice.

| Chr | Position_hg19 | Nearby gene | SNP ID | Effect allele | Other allele | Effect allele frequency | Statin |  | Fibrate |  | Ezetimibe |  | FinnGen* |  |
| --- | --- | --- | --- | --- | --- | --- | --- | --- | --- | --- | --- | --- | --- | --- |
|  |  |  |  |  |  |  | log_OR | P | log_OR | P | log_OR | P | P | Other associated traits** |
| 1 | 55,505,647 | *PCSK9* | rs11591147 | G | T | 0.99 | **0.239** | **4.9E-09** | -0.147 | 4.3E-01 | 0.731 | 1.6E-06 | **0** | LDL |
| 1 | 109,818,306 | *PSRC1* | rs629301 | G | T | 0.20 | **-0.063** | **3.0E-08** | -0.101 | 6.0E-02 | -0.182 | 5.5E-07 | **3.0E-174** | LDL |
| 2 | 21,266,774 | *APOB* | 2:21266774_ | GGCAGCGCCA | G | 0.64 | -0.026 | 6.6E-03 | -0.046 | 3.1E-01 | **-0.170** | **3.5E-09** | NA | LDL |
| 2 | 27,730,940 | *GCKR* | rs1260326 | T | C | 0.40 | **0.051** | **2.6E-08** | 0.220 | 3.0E-07 | 0.078 | 6.4E-03 | **6.2E-36** | TG |
| 6 | 161,005,610 | *LPA* | rs55730499 | C | T | 0.91 | **-0.104** | **5.4E-11** | -0.040 | 6.0E-01 | **-0.332** | **1.7E-13** | **8.1E-82** | Lipoprotein(a), LDL |
| 6 | 161,089,307 | *LPA* | rs56393506 | C | T | 0.82 | **-0.089** | **1.1E-13** | 0.076 | 1.9E-01 | -0.172 | 1.8E-06 | **4.3E-53** | Lipoprotein(a), LDL |
| 8 | 9,184,231 | *RP11-10A14.4* | rs4240624 | G | A | 0.09 | **-0.097** | **6.8E-10** | -0.010 | 8.9E-01 | -0.042 | 3.9E-01 | **1.2E-26** | HDL |
| 9 | 22,103,813 | *CDKN2B-AS1* | rs1333042 | A | G | 0.50 | **-0.081** | **6.1E-19** | -0.075 | 8.4E-02 | -0.050 | 7.8E-02 | **3.3E-42** | CAD |
| 9 | 107,661,742 | *ABCA1* | rs2740488 | A | C | 0.74 | **0.073** | **1.7E-12** | 0.036 | 4.6E-01 | 0.079 | 1.3E-02 | **6.7E-16** | HDL |
| 10 | 114,754,784 | *TCF7L2* | rs35198068 | T | C | 0.70 | **-0.066** | **2.5E-11** | -0.001 | 9.8E-01 | -0.034 | 2.7E-01 | **5.0E-19** | T2D |
| 11 | 61,593,816 | *FADS2* | rs174568 | C | T | 0.66 | 0.030 | 1.6E-03 | **-0.244** | **2.8E-08** | -0.019 | 5.3E-01 | **5.0E-39** | TG |
| 11 | 116,648,917 | *ZNF259* | rs964184 | G | C | 0.15 | 0.012 | 3.5E-01 | **0.622** | **2.2E-34** | 0.199 | 1.1E-07 | **3.5E-155** | TG, HDL |
| 15 | 58,679,807 | *LIPC* | 15:58679807 | CAGA | C | 0.65 | **-0.093** | **3.1E-22** | -0.122 | 6.0E-03 | -0.084 | 4.2E-03 | NA | HDL |
| 15 | 91,420,973 | *FURIN* | rs1573643 | T | C | 0.67 | **-0.061** | **2.5E-10** | -0.027 | 5.6E-01 | -0.005 | 8.6E-01 | 1.5E-05 | Blood pressure |
| 16 | 57,006,590 | *CETP* | rs7499892 | C | T | 0.80 | **0.160** | **2.8E-44** | 0.078 | 1.5E-01 | 0.060 | 9.1E-02 | **1.8E-09** | HDL |
| 16 | 75,456,304 | *CFDP1* | rs12934767 | T | C | 0.42 | **-0.052** | **1.7E-08** | -0.055 | 2.2E-01 | 0.013 | 6.5E-01 | NA | CAD |
| 19 | 45,413,233 | *APOE* | rs1065853 | G | T | 0.94 | 0.030 | 1.1E-01 | **-0.583** | **4.5E-16** | 0.260 | 3.0E-05 | **0** | LDL, HDL, TG |

The variant with smallest P in each locus is shown for each drug. Genome-wide significant association (P <5e-8) in bold.
\*Statin medication use GWAS in FinnGen.
\*\*Looked up in https://hugeamp.org/ and https://atlas.ctglab.nl/PheWAS

Among UK Biobank EUR hypertensive subjects (Table 2B), we identified genome-wide significant loci for drug choice: 6 for diuretic, 3 for β-blocker, 8 for calcium channel blocker, and 6 for RAS-acting agents (Table 4). These loci also overlapped with those associated with blood pressure, lipid levels and CAD, indicating shared genetic architecture across cardiometabolic traits. Comparison with antihypertensive *use* GWAS in FinnGen showed replication for 16 of 18 SNPs (89%) at genome-wide significance.

**Table 4.**
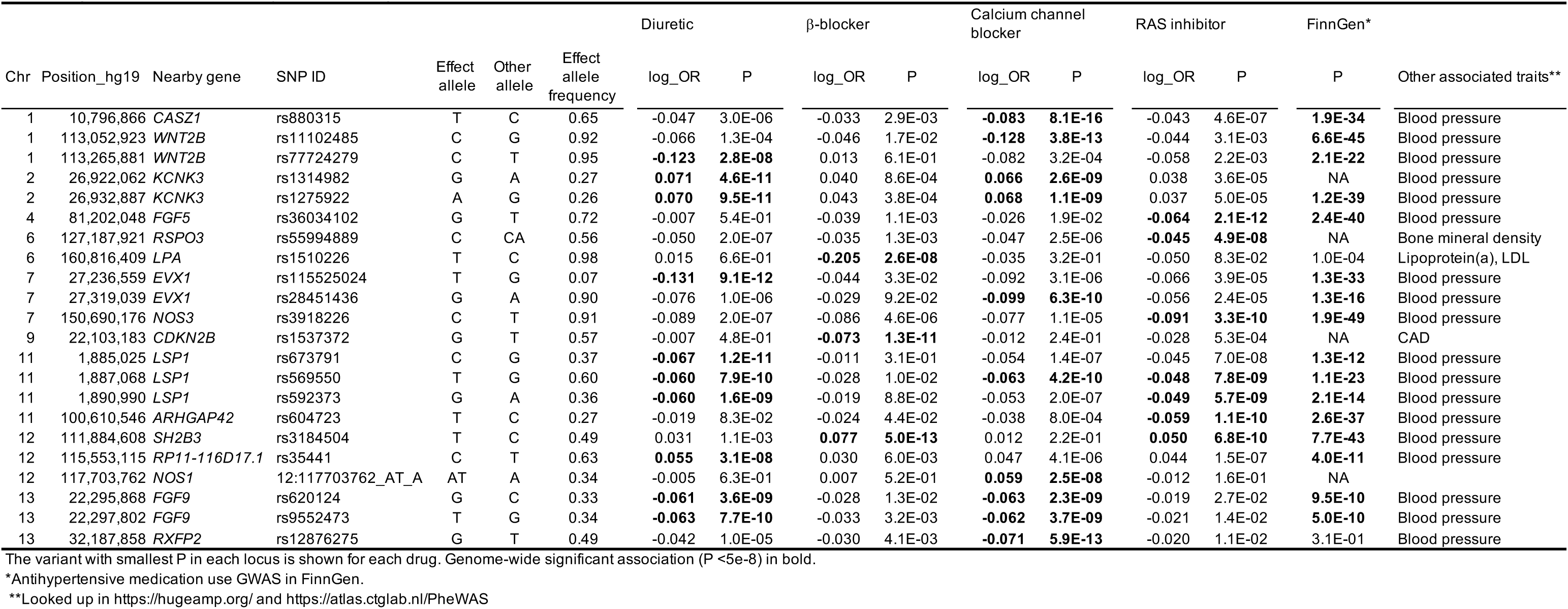
Significant loci for GWAS of hypertension drug choice.

### 3.4. Genetic correlations between drug choice and disease traits

To characterize the shared genetic basis between drug choice and cardiometabolic diseases, we estimated SNP heritability and genetic correlations (Figure 2). Among dyslipidemia medications, statin showed the highest heritability (0.054, SE = 0.005), followed by ezetimibe (0.011, SE = 0.003) and fibrate (0.008, SE = 0.004). Statin choice correlated most strongly with blood pressure and CAD, fibrate choice with HDL-C (negative direction) and triglycerides, and ezetimibe choice with LDL-C. All three medications showed moderate correlations with T2D.

**Figure 2.**
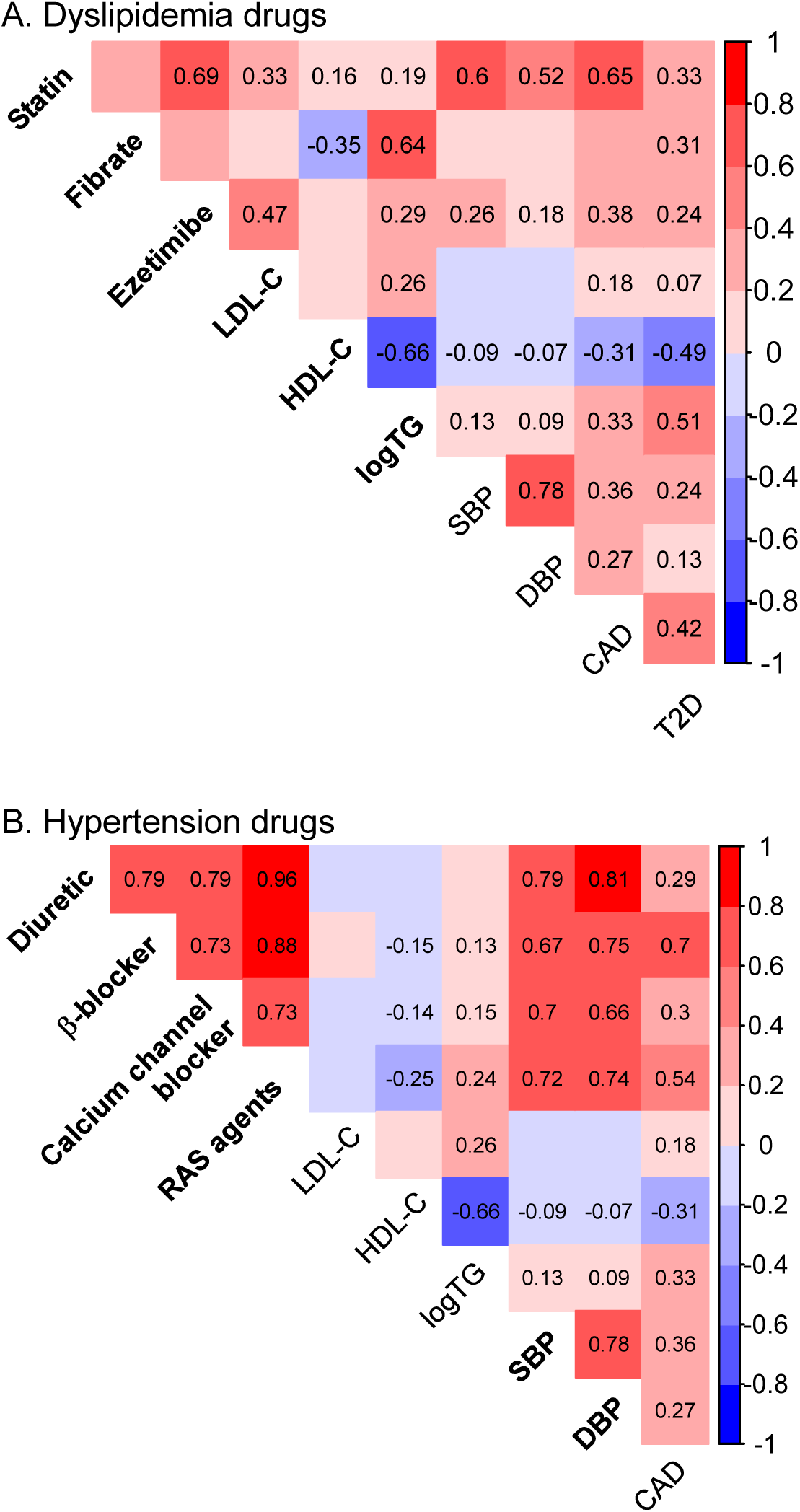
Genetic correlation analysis. Heatmaps shows genetic correlations between drug choice traits and cardiometabolic disease traits for dyslipidemia (**A**) and hypertension (**B**) medications. Disease traits include LDL-C, HDL-C, triglyceride levels (logTG), systolic blood pressure (SBP), diastolic blood pressure (DBP), coronary artery disease (CAD) and type 2 diabetes (T2D). Color intensity indicates correlation magnitude, and numerical values are displayed for significant correlations (P-value <0.01). Drug names and their primary target disease traits are highlighted in bold font.

For hypertension medications, RAS-acting agents had the highest heritability (0.033, SE = 0.004), followed by calcium channel blocker (0.030, SE = 0.004), diuretic (0.019, SE = 0.004), and β-blocker (0.017, SE = 0.003) for. All antihypertensives displayed mutual genetic correlations and significant correlations with blood pressure. β-blocker choice correlated most strongly with CAD, while RAS-acting agents correlated most strongly with HDL-C (negative direction) and triglycerides.

### 3.5. PGS for drug choice

We developed PGS to predict drug choice, leveraging the observed genetic overlap between drug choice and cardiometabolic traits (Tables 3, 4, and Figure 2). A total of 570 SNPs associated with relevant traits from published literature were compiled and weighted to generate predictive models. The model development utilized 80% of UK Biobank EUR subjects, with validation performed on the remaining UK Biobank subjects, the All of Us cohort, and the UK Biobank South Asian descent (SAS) subjects (Table 2). Alongside the PGS, we derived a basic score based on sex, age and BMI, as well as a combined score by summing the basic score and PGS.

For dyslipidemia drugs, validation in UK Biobank EUR showed varying predictive performance measured by the Area Under the Receiver Operating Characteristic Curve (AUC-ROC): 0.550–0.621 for the PGS, 0.619–0.694 for the basic score, and 0.633–0.702 for the combined score (Table 5A). Although the combined score demonstrated a statistically significant improvement over the basic score, the increase was modest (0.008–0.056). In the All of Us cohort, PGS showed modest yet statistically significant performance. The AUC-ROC for PGS was similar or larger in UK Biobank SAS subjects than in the EUR subjects, even though the scores were trained in EUR subjects. Among the three drugs, fibrate achieved the highest prediction performance across all cohorts.

**Table 5A.** Prediction of dyslipidemia drug choice.

| Drug | AUC-ROC in UK Biobank, EUR |  |  |  | AUC-ROC in All of Us, EUR |  | AUC-ROC in UK Biobank, SAS |  |
| --- | --- | --- | --- | --- | --- | --- | --- | --- |
|  | PGS | Basic | Combined | P, AUC<sub>basic</sub> < AUC<sub>combined</sub> * | PGS | P, AUC<sub>PGS</sub> >0.5 * | PGS | P, AUC<sub>PGS</sub> >0.5 * |
| Statin | 0.555 | 0.694 | 0.702 | 1.5E-17 | 0.516 | 6.6E-16 | 0.572 | 1.9E-13 |
| Fibrate | 0.621 | 0.619 | 0.675 | 4.3E-08 | 0.614 | 1.3E-121 | 0.620 | 4.1E-03 |
| Ezetimibe | 0.550 | 0.622 | 0.633 | 4.0E-03 | 0.549 | 7.3E-27 | 0.554 | 5.5E-02 |

Analysis of drug choice frequencies by score deciles in UK Biobank EUR (Figure 3A) revealed clear gradients across both the PGS and basic score, with the combined score showing the greatest separation. In the top decile for the combined statin score, sensitivity—the proportion of individuals prescribed the drug—was 71%. Similar patterns were observed in the All of Us cohort and UK Biobank SAS, though gradients in UK Biobank SAS were less uniform due to the smaller sample size. Notably, drug-specific PGS were more predictive for their intended drugs than PGS for other medications, confirming their specificity (Supplementary Figure 2A). Statin prescriptions increased with higher statin PGS (first row) and a similar trend was observed for fibrate (second row). However, ezetimibe PGS did not outperform other PGS in predicting ezetimibe prescription.

**Figure 3.**
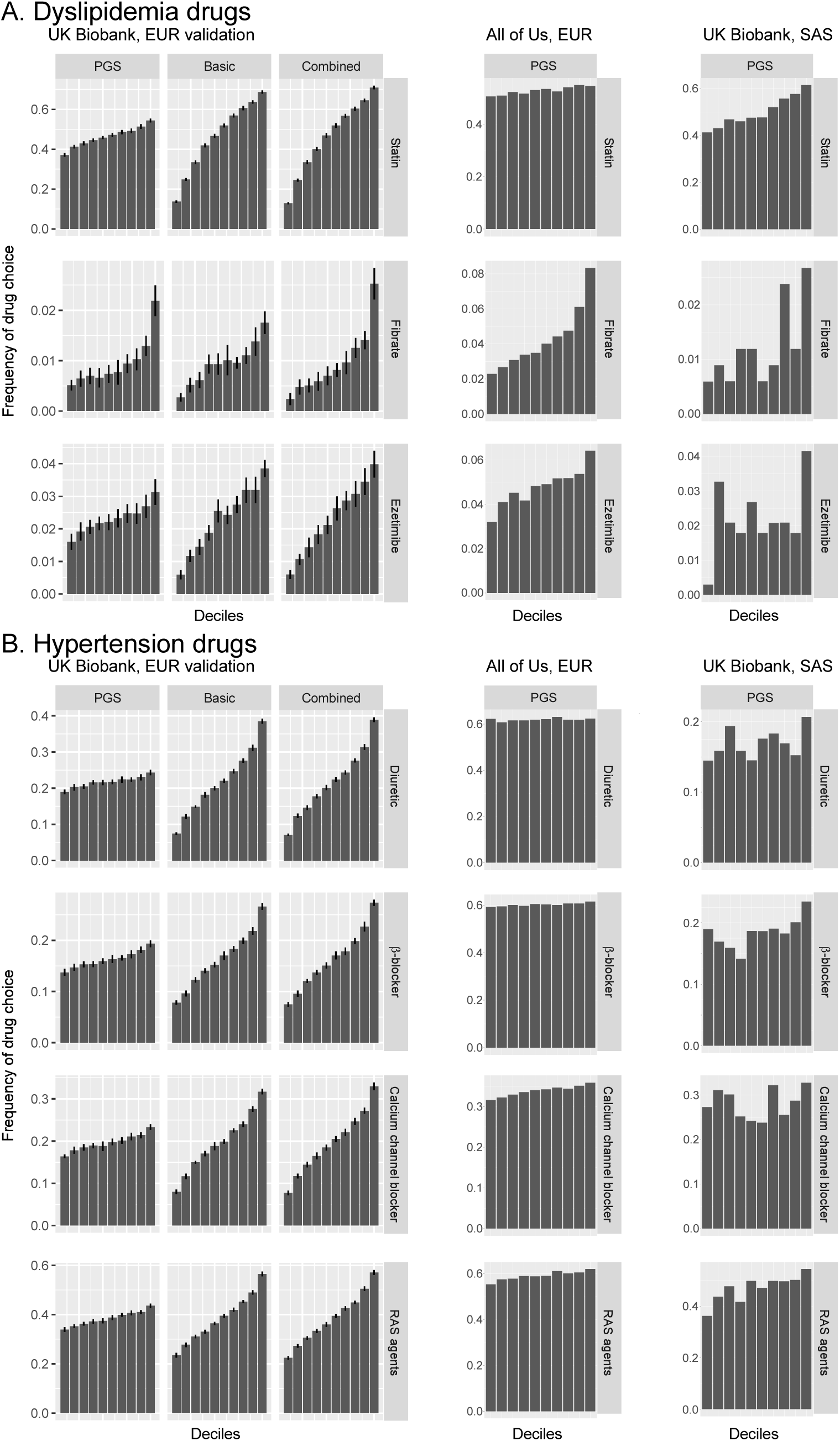
Drug choice frequencies across PGS deciles. This figure compares drug choice frequencies across score deciles for dyslipidemia (**A**) and hypertension (**B**) medications in the UK Biobank EUR validation cohort (left panels), the All of Us cohort (middle panels), and UK Biobank SAS (right panels). Each row corresponds to a specific drug, and columns represent different prediction score types (PGS, basic score, and combined score). Bar heights indicate the proportion of subjects prescribed the drug within a given decile. In UK Biobank EUR, error bars represent the standard deviation across 20 cross-validation trials. EUR, European descent; SAS, South Asian descent.

For hypertension drugs, overall predictive performance was lower. In UK Biobank EUR, AUC-ROC values were 0.524–0.534 for PGS, 0.614–0.649 for the basic score, and 0.620–0.651 for the combined score (Table 5B). Although the improvements from adding PGS to the basic score were statistically significant, they remained small (0.002–0.007). In the All of Us cohort, the PGS achieved statistical significance for all antihypertensives except diuretic. In UK Biobank SAS, AUC-ROC values were comparable to those of UK Biobank EUR.

**Table 5B.** Prediction of hypertension drug choice.

| Drug | AUC-ROC in UK Biobank, EUR |  |  |  | AUC-ROC in All of Us, EUR |  | AUC-ROC in UK Biobank, SAS |  |
| --- | --- | --- | --- | --- | --- | --- | --- | --- |
|  | PGS | Basic | Combined | P, AUC<sub>basic</sub> < AUC<sub>combined</sub> * | PGS | P, AUC<sub>PGS</sub> >0.5 * | PGS | P, AUC<sub>PGS</sub> >0.5 * |
| Diuretic | 0.524 | 0.649 | 0.651 | 1.2E-04 | 0.503 | 0.07 | 0.517 | 0.11 |
| β-blocker | 0.533 | 0.614 | 0.620 | 1.1E-06 | 0.507 | 6.0E-04 | 0.532 | 1.1E-02 |
| Calcium channel blocker | 0.534 | 0.624 | 0.630 | 4.8E-08 | 0.516 | 4.3E-14 | 0.508 | 0.25 |
| RAS-acting agents | 0.534 | 0.617 | 0.624 | 5.5E-11 | 0.521 | 4.6E-24 | 0.549 | 2.7E-06 |

EUR, European descent; SAS, South Asian descent; \*alternative hypothesis

Decile analysis in UK Biobank EUR demonstrated modest PGS gradients for hypertension drugs but more pronounced gradients for the basic score, with marginal gains from combining the two (Figure 3B). For the top decile of combined score for RAS-acting agents, the sensitivity was 57%. In the All of Us cohort and UK Biobank SAS, PGS showed the clearest separation for RAS-acting agents. Drug-specific PGS demonstrated stronger predictive power for their intended medications than PGS for other medications, although this specificity was less pronounced than for dyslipidemia drugs (Supplementary Figure 2B).

## 4. Discussion

We investigated whether germline genetic variation influences drug response and drug choice for dyslipidemia and hypertension, using data from the UK Biobank and All of Us cohorts. Although prior studies reported SNPs modifying LDL-C response to statins, we found that these associations disappeared once the biomarker scaling issue was addressed. In contrast, GWAS for drug choice identified several significant loci overlapping with established cardiometabolic disease loci. Genetic correlation analysis revealed distinct patterns of genetic sharing between drug choice and disease traits. Building on these observations, we developed drug choice PGS using SNPs associated with relevant cardiometabolic traits. The PGS demonstrated modest yet statistically significant predictive performance, especially for dyslipidemia medication choice (AUC-ROC 0.550–0.621 in UK Biobank). When combined with demographic factors (age, sex and BMI), predictive performance increased further (AUC-ROC 0.633–0.702). External validation in the All of Us cohort confirmed the generalizability of these findings, although with slightly attenuated performances. Notably, the PGS developed in UK Biobank EUR performed comparably in UK Biobank SAS, suggesting cross-ancestry transferability.

### 4.1. Spurious drug response associations caused by biomarker scaling

Pharmacogenetic studies typically quantify drug-induced biomarker response as either absolute change (on-drug level *Y* minus baseline level *X*) or relative change, computed as *Y*/*X* – 1 or log(*Y*/*X*), which are mutually convertible and yield similar values. Previous GWAS of LDL-C response to statins have used measures of absolute change [29,30] or relative change [2,29–32].

Our analysis revealed that genetic variants associated with baseline LDL-C display systematically opposite associations with absolute LDL-C change (Figure 1B, left panel) but concordant associations with relative change (Figure 1B, right panel) in response to statin therapy. Absolute change represents the difference between raw LDL-C values (*Y* – *X*), while relative change represents the difference between log-transformed values (log(*Y*) – log(*X*)). After applying a variance-stabilizing Box-Cox transformation, which represents an intermediate scaling between identity and logarithmic transformations and equalizes drug effects across the baseline biomarker range (Figure 1C bottom panel, 1D bottom panel), no variants remained significantly associated with LDL-C response to statins.

Previous studies recognized that variants associated with baseline LDL-C can spuriously appear associated with LDL-C change, leading some researchers to adopt baseline-adjusted residuals as the target phenotype [33]. However, introduction of false associations by baseline adjustment has been demonstrated through analytical derivations [29,34], simulation studies [35], and empirical analyses[29,32]. A previous study[33] employed the Box-Cox transformation with a different objective, to ensure that the residuals from the regression of on-drug LDL-C level on baseline LDL-C fits a Gaussian distribution. Here the optimal parameters for power transformation were λ = 0.16, 0.48 and 0.68 in three cohorts, values lying between the identity and logarithmic transformations.

To our knowledge, this study is the first to demonstrate that biomarker scaling methods can systematically generate spurious pharmacogenetic signals and to propose the variance-stabilizing Box-Cox transformation as a solution. This methodological insight has broad implications for pharmacogenomic research, suggesting that some previously reported genetic associations with drug response may be artifacts arising from skewed data distributions—particularly when the optimal Box-Cox parameter substantially differs from the identity (λ = 1) or logarithmic (λ = 0) cases. Future pharmacogenetic studies should consider applying variance-stabilizing transformations to mitigate these artifacts and to ensure more reliable identification of genuine genetic predictors of drug response.

### 4.2. Genetic determinants of drug choice trait

This study introduces the concept of genetic determinants of the “drug choice trait”, demonstrated through development of PGS for dyslipidemia and hypertension medication choices. Our approach uniquely defines cases as patients prescribed a given drug and controls as those with the same disease who were not prescribed that drug. This differs from previous studies on the “drug use trait,” which defined controls as all non-drug users, including healthy individuals [6]. In the context of personalized medicine, the former trait captures which drug to pick at the time of diagnosis, and the latter trait captures the drug to pick regardless of disease onset. For example, statin drug choice in UK Biobank and statin drug use in FinnGen [18] showed a moderate genetic correlation of 0.644 (SE: 0.005).

The genetic architecture of drug choice traits showed varying degrees of overlap with cardiometabolic disease traits (Figure 2), consistent with known pharmacological mechanisms. Among dyslipidemia medications, fibrate choice correlated most strongly with triglycerides, consistent with its PPARα-activation mechanism and effects on fatty acid catabolism. Ezetimibe choice correlated primarily with LDL-C, reflecting its clinical role as an add-on therapy when statin monotherapy is insufficient. Statin choice correlated most strongly with blood pressure and CAD, aligning with its widespread use in cardiovascular risk management. For hypertension medications, β-blocker choice demonstrated the strongest correlation with CAD, in line with its mechanism of reducing catecholamine effects and heart rate. The correlation between RAS-acting agent choice and triglycerides corresponds with RAS’s known influence on hepatic and skeletal muscle fatty acid metabolism [36].

### 4.3. Limited utility of drug choice PGS

Although germline genetics may eventually guide medication choice, our current PGS for hypolipidemic and antihypertensive drug choice offers limited predictive utility. In this study, individuals in the top combined-score decile exhibited 71% sensitivity for statin and 57% for RAS-acting agent prescriptions, but lower sensitivity for other drug classes (Figure 3).

Our PGS methodology builds upon the concept of partitioned PGS for diabetes [37,38], but instead of partitioning SNPs into subgroups, we weighted cardiometabolic disease SNPs based on their association with drug choice traits. This approach differs from previous pharmacogenomic PGS studies, which focused on predicting drug outcomes (toxicity, efficacy, or optimal dosage) either through small drug trial cohorts [39] or by leveraging disease-related PGS [40]. Appropriation of disease-related PGS for drug choice prediction showed variable performance across different drug classes in our analyses. For the antihypertensives, which exhibit high genetic correlation with blood pressure (Figure 2B), the drug choice traits could be predicted by the PGS for hypertension (AUC-ROC 0.545–0.561) slightly more accurately than by the PGS’s trained for the respective drugs. In contrast, ezetimibe, statin and fibrate exhibit moderate to no genetic correlation with LDL-C (Figure 2A), and the LDL-based PGS showed performance ranging from aligned to inverse direction (AUC-ROC 0.584, 0.517 and 0.487, respectively).

Drug suitability can be conceptualized as the composite of safety, efficacy, and pathological appropriateness. Simplistically, interindividual variability in drug suitability can be regarded as a combination of interindividual variability in the three factors. For cardiometabolic disease drugs, the variability in safety is likely minimal because widely used drugs generally lack severe toxicity in subpopulations. In terms of efficacy, known genetic variants affecting drug efficacy reportedly show relatively small effects (e.g., ∼1% effect on LDL-C by variants in *LPA*, *APOE*, *SORT1*, *SLCO1B1* for statins, and odds ratio of 1.25 for metformin response in *ATM* variants) [1] although our analysis suggests some reported effects may reflect methodological artifacts. Consequently, genetic difference in drug suitability may predominantly arise from interindividual variability in pathological appropriateness.

Although our drug choice PGS exhibited statistically significant predictability, the modest AUC-ROC values indicate room for improvement. Enhancements could stem from expanding the SNP set and refining allele weighting. Our current PGS includes 570 SNPs derived from cardiometabolic disease GWAS published until 2017, deliberately excluding UK Biobank and All of Us data to ensure rigorous validation. Incorporating more recent large-scale GWAS findings and anticipated studies of rare variants with greater effect sizes would expand the SNP set. More importantly, refining SNP allele weighting with data from large genomic-prescription cohorts is necessary to increase accuracy.

While drug choice PGS could eventually assist in clinical medication matching, several limitations remain. First, the PGS captures real-world prescribing behavior rather than direct pharmacodynamic or outcome-based efficacy. A higher PGS does not necessarily imply a superior therapeutic response, which can only be evaluated through randomized clinical trials. Although clinical trial-based pharmacogenomic studies enable precise assessment of drug dose and administration, strict trial inclusion criteria may introduce selection bias. Second, we did not analyze how cross-cohort difference in clinical guidelines, such as those between the NICE versus American recommendations and their temporal versions, influence medication choice in UK Biobank and All of Us. Third, prescribed medications in the training data may not always represent optimal patient-drug matches. The modest predictive performance of genetics alone underscores the need for integrated models combining genetic, demographic and clinical features to achieve more practical drug recommendation systems.

## Ethical declaration

The UK Biobank is approved by the North West Multi-centre Research Ethics Committee as a Research Tissue Bank. The All of Us Research Program is approved and managed through a dedicated Institutional Review Board. This research was conducted in accordance with the Declaration of Helsinki and all relevant institutional and national guidelines for research involving human participants.

## Data Availability

Code files for analyses in this manuscript are available from https://github.com/fumi-github/drug_selection

## Acknowledgements

This research was conducted using the UK Biobank Resource under Application Number 55469. We gratefully acknowledge All of Us participants for their contributions, without whom this research would not have been possible. We also thank the National Institutes of Health’s All of Us Research Program for making available the participant data examined in this study. We also thank the participants and investigators of the FinnGen study.

## Author contributions

Conceptualization, Formal analysis, Writing – Original Draft: F.T.; Writing – Review & Editing: F.T., M.S.I.D., W.W.H.H., S.A.L., M.I. and N.K.

---

**Supplementary Figure 1.**
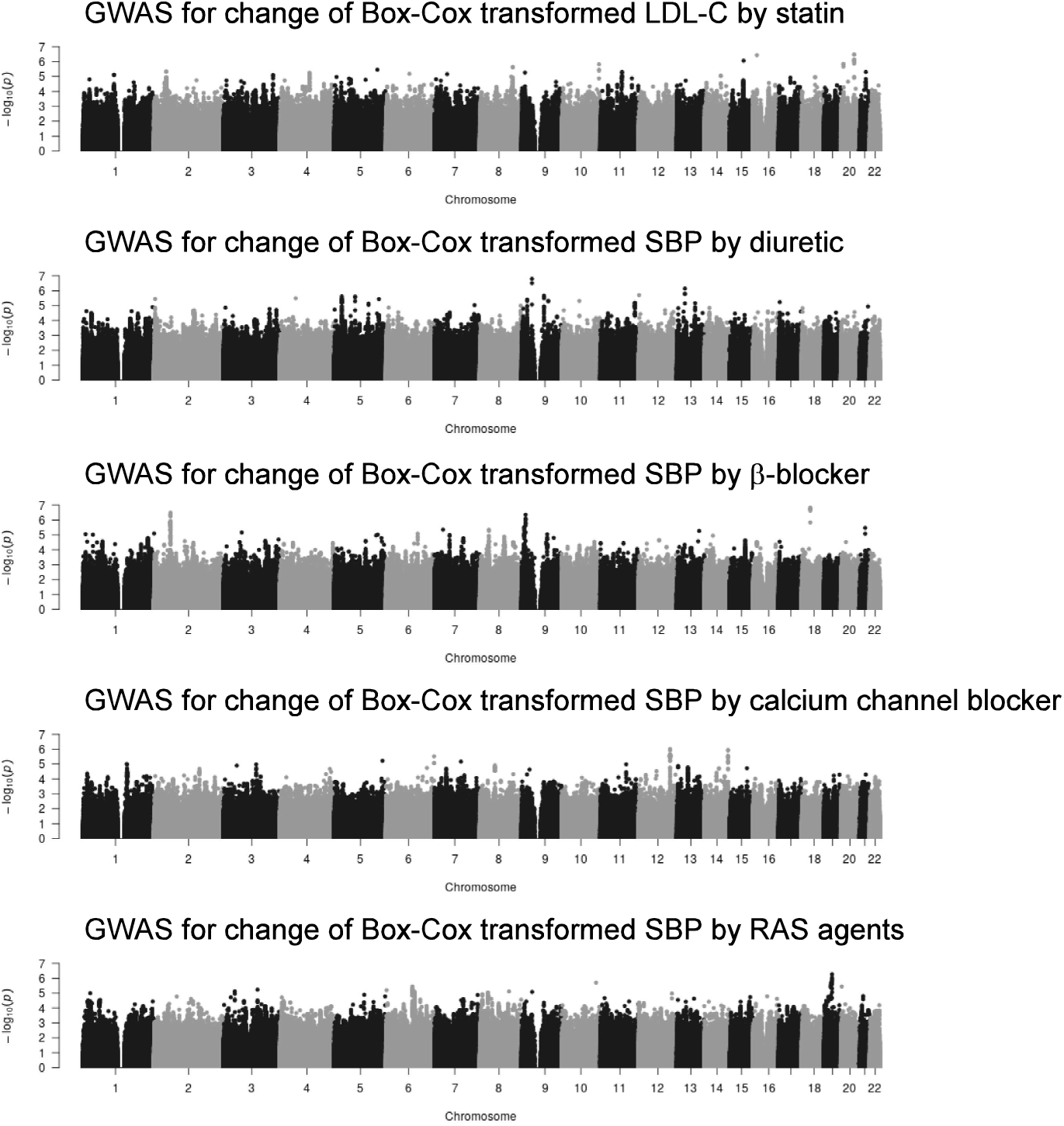
Manhattan plots for GWAS of drug-induced biomarker response after variance-stabilizing Box-Cox transformation. Variants are plotted as points, with chromosomal position along the horizontal axis, and –log_10_(P-value) along the vertical axis.

**Supplementary Figure 2.**
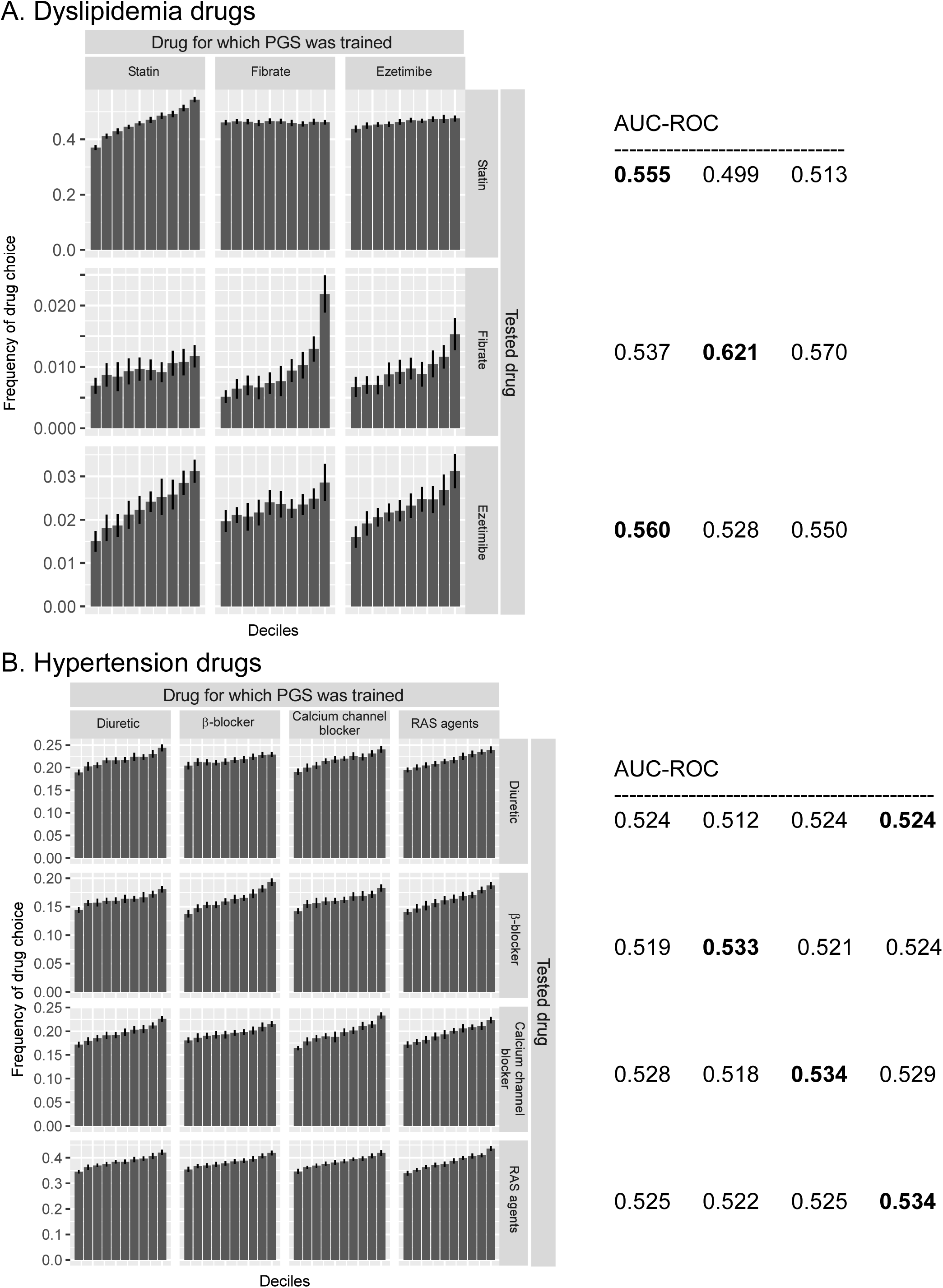
Performance of drug-specific PGS for dyslipidemia (**A**) and hypertension (**B**). Left panels illustrate cross-comparisons of drug choice frequencies across PGS deciles trained for different medications in the UK Biobank EUR subjects. Rows represent the tested drugs, and columns represent the drugs used for PGS training. Bar heights indicate the proportion of subjects prescribed each drug across PGS deciles, with error bars indicating the standard deviation from cross-validation trials. The tables on the right summarize the corresponding Area Under the Receiver Operating Characteristic Curve (AUC-ROC) for each combination. For each tested drug (row), the model with the highest AUC-ROC is highlighted in bold font. This analysis demonstrates the specificity of each drug’s PGS for predicting its own intended prescription pattern.

